# Rethinking the Impact of Pretransplant Malignancy (Pre-TM) on Double Lung Transplantation (DLT) Eligibility: An Analysis of 23,291 DLT Recipients

**DOI:** 10.1101/2024.03.14.24304302

**Authors:** Wongi Woo, Hye Sung Kim, Ankit Bharat, Young Kwang Chae

## Abstract

**Background:** Given the increasing need for lung transplants among older patients with a history of cancer, this study analyzed database registry to assess outcomes for DLT recipients with Pre-TM.

**Methods:** This study evaluated the United Network for Organ Sharing (UNOS) registry for adult DLT performed between 2005 and 2023. Patients with a history of previous or multi-organ transplants, and those with donors who had cancer history, were excluded. Propensity-score matching was used to compare patients with or without Pre-TM. Overall and Post-TM-free survival were analyzed.

**Results:** Among the 23,291 recipients of DLT, 8.0%(1,870) had Pre-TM. Compared to those without Pre-TM, patients with Pre-TM had worse overall (hazard ratio[HR] 1.20, 95% confidence interval[CI] 1.12-1.29, p<0.001) and Post-TM-free survival (HR 1.32, 95% CI 1.24-1.41, p<0.001). However, after adjusting for age, sex, and race through propensity-score matching, the survival difference between the groups became non-significant (HR 1.05, 95% CI 0.97-1.13, p=0.229). While the Pre-TM group still had worse Post-TM-free survival, this difference diminished after excluding cutaneous Post-TM (HR 1.06, 95% CI 0.99-1.15, p=0.116). Additionally, the recurrence rate of Pre-TM after transplant wasn’t higher than de novo cancers in patients without Pre-TM.

**Conclusion:** Patients with Pre-TM had similar overall survival rates after DLT as those without Pre-TM. Importantly, there is no increased risk of the primary Pre-TM type recurring post-transplant compared to patients without Pre-TM. These findings highlight the necessity for a more nuanced evaluation of transplant candidacy to prevent premature exclusion of Pre-TM patients from life-saving surgeries.

## Introduction

The clinical outcomes of lung transplants have markedly improved since they were first performed in 1963. Data from the International Society for Heart and Lung Transplantation (ISHLT) reveal a dramatic increase in double lung transplant (DLT) recipients, from 69 in 1988 to 4,452 in 2017^1^. This impressive progress is due to a synergy of factors: the standardization of surgical techniques^2^, the implementation of more effective allocation systems^3^, refinements in immunosuppressive therapy^4^, and the introduction of innovative technologies, such as Ex-Vivo lung perfusion.^5^

As lung transplantation has gained widespread acceptance, new challenges have emerged. The re-evaluation of previously established exclusion criteria, such as age, viral infections (including HIV, hepatitis, and COVID-19), and cardiovascular comorbidities, reflects a progression towards more inclusive practices. One of the most pressing issues is the rise in older patients with multiple comorbidities, especially those with a malignancy history. Historically, any history of malignancy was deemed an absolute contraindication for lung transplantation due to the elevated risk of post-transplant malignancy (Post-TM),^6^ a concern compounded by the intensive immunosuppression needed to prevent organ rejection.^7^ Nevertheless, the stance on this issue has evolved. The 2021 ISHLT consensus has begun to acknowledge the potential eligibility of patients with pre-transplant malignancy (Pre-TM) for transplantation^8^, although the lack of robust evidence supporting these guidelines underscores ongoing uncertainty.

Analyses of United Network for Organ Sharing (UNOS) datasets have taken various forms to explore the impact of cancer history on the clinical outcomes of transplant recipients. These studies have found Pre-TM to be a significant risk factor for overall survival and the development of malignancies post-transplant, both in kidney and lung transplant recipients.^6,9^ Despite these insights, interpreting these results faces certain obstacles. First, distinctions were not made between DLT and single lung transplants, which are known to have different prognoses. Secondly, demographic and genetic factors, such as sex and race, proven to be significant in determining prognoses in extensive database studies, were not considered in the analysis.^10,11^ In addition, there is a need to update this research with more recent data, particularly in light of advances in cancer treatment like immunotherapy and targeted therapy. Finally, a notable gap exists in research comparing the post-transplant recurrence rates of Pre-TM with the incidence rates of de novo Post-TM of the same cancer type.

This study aims to dissect the clinical outcomes for DLT recipients in the contemporary era, with a particular focus on individuals with Pre-TM, while adjusting for critical demographic factors.

## Methods

### Study population and outcome of interest

This study retrospectively reviewed data from adult patients who underwent DLT in the UNOS registry from January 2005 to September 2023. Patients with a history of previous or multi-organ transplants, and those with donors who had a history of malignancy, were excluded. Pre- and post-transplant malignancy types were classified into twelve categories based on the organ system, according to National Cancer Institute guidelines: breast, digestive, genitourinary, gynecologic, head and neck, hematologic, musculoskeletal, neurologic, respiratory, skin, unknown primary, and others. The codes for Pre- and Post-TM were reviewed by two authors, and any disagreement in classification was resolved by a third author. Patients with two major cancer types were classified as having multiple cancers.

The primary outcome was overall survival, and the secondary outcome was Post-TM-free survival. Post-TM-free survival was defined as the time from DLT to the occurrence of Post-TM or death from any cause.

### Pre-TM and Post-TM clinical outcomes analysis

Subgroup analysis was performed to evaluate the post-transplant recurrence rate of the original cancer in patients with Pre-TM. Given the lack of time-specific data, comparisons of recurrence and incidence rates were conducted using Chi-square analysis. For example, the post-transplant recurrence rate of breast cancer in patients with a history of pre-transplant breast cancer was compared with the incidence rate of post-transplant breast cancer in patients without Pre-TM. This comparative method was applied across seven different cancer types to assess recurrence patterns.

### Statistical analysis

Continuous variables were presented as medians and interquartile ranges (IQR), and the t-test was used for their analysis. The Chi-square test was used to compare categorical variables. Cox proportional hazard analysis identified relevant risk factors for overall-survival and Post-TM-free survival. The Kaplan–Meier log-rank test compared the clinical outcomes of patients with or without Pre-TM. To adjust for unbalanced confounding variables, propensity-score matching compared these two groups. The propensity score for each participant was calculated using a logistic model that included age, sex, and race. Subsequently, nearest-neighbor matching within 0.2 caliper width without replacement facilitated 3:1 matching of patients between the two groups. Statistical analyses were performed using R version 4.0.4 (R Core Team, R Foundation for Statistical Computing, Vienna, Austria), with differences considered statistically significant at a two-tailed p-value of□<□0.05.

## Results

### Characteristics of the study cohort

Among the 32,562 patients who underwent lung transplants during the study period, 23,291 (71.5%) patients received DLT. The median age was 58.0 years (IQR 48-64), and 58.3% (13,582/23,291) were male (Table 1). Approximately half of the patients had a smoking history (55.4%, 12,708/22,938), and the majority had worse a performance scale, predominantly ECOG 2 or 3.

### Pre-transplant malignancy

A total of 8.0% (1,870/23,291) had a history of cancer before undergoing DLT (Table 2). Skin cancer constituted the largest proportion at 39.0% (726/1,870), followed by breast (10.6%, 197/1,870), genitourinary (10.3%, 191/1,870), hematologic (7.9%, 148/1,870). A significant portion

(20.1%, 374/1870) had no information regarding their specific Pre-TM diagnosis, and 6.1% (113/1870) of the patients had histories of multiple cancers.

When compared with patients without Pre-TM, the Pre-TM group was older, had a higher proportion of males, and was predominantly Caucasian (Table 1). Regarding indications for DLT, the Pre-TM group had more patients with COPD and IPF, and fewer with CF. These disparities became less pronounced following propensity-score matching (Table 2).

Overall, patients with Pre-TM had worse clinical outcomes in terms of overall survival (p<0.001, Figure 1A). However, the difference was not observed after propensity score matching (p=0.226, Figure 1B).

**Figure 1.**
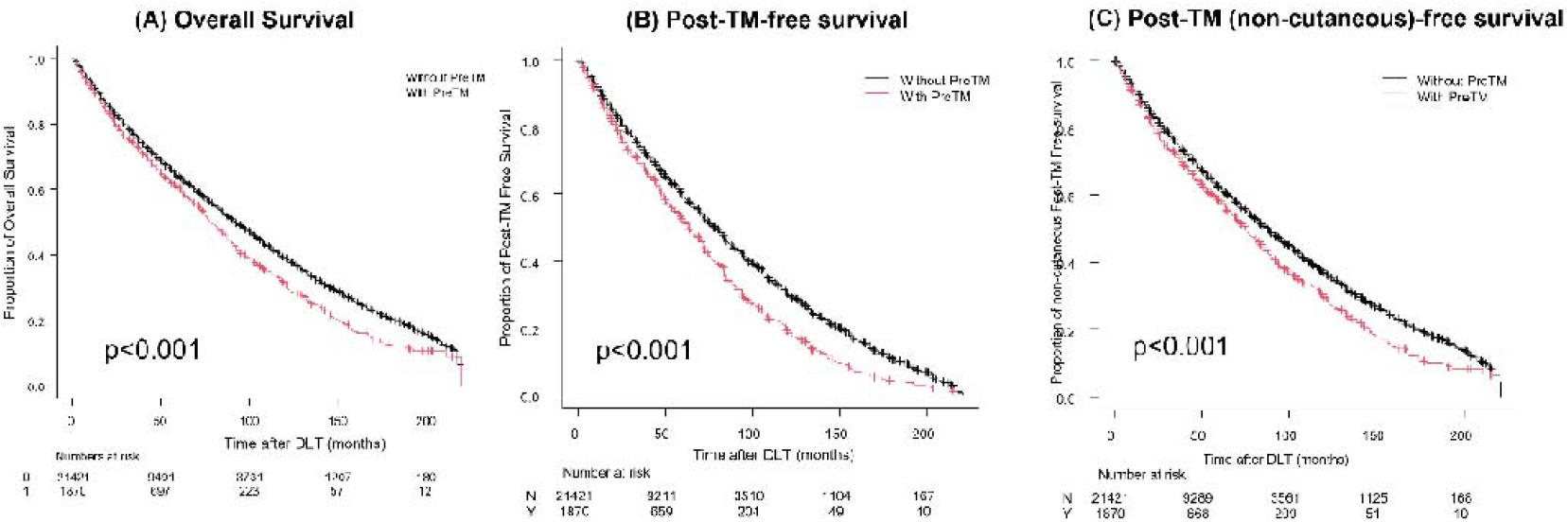
Clinical outcomes of the entire cohort according to the status of pre-transplant malignancy. Pre-TM, pre-transplant malignancy; Post-TM, post-transplant malignancy

### Post-transplant malignancy

The incidence of Post-TM among all patients was 19.8% (4,623/23,291), and it was higher among those with Pre-TM (Supplementary Table S1). A similar pattern persisted even after propensity-score matching for patients with Pre-TM. In terms of Post-TM-free survival, the Pre-TM group exhibited poorer clinical outcomes (Figures 2A and 2B). Yet, when cutaneous malignancies were excluded, this difference was not observed after propensity-score matching (Figure 3).

**Figure 2.**
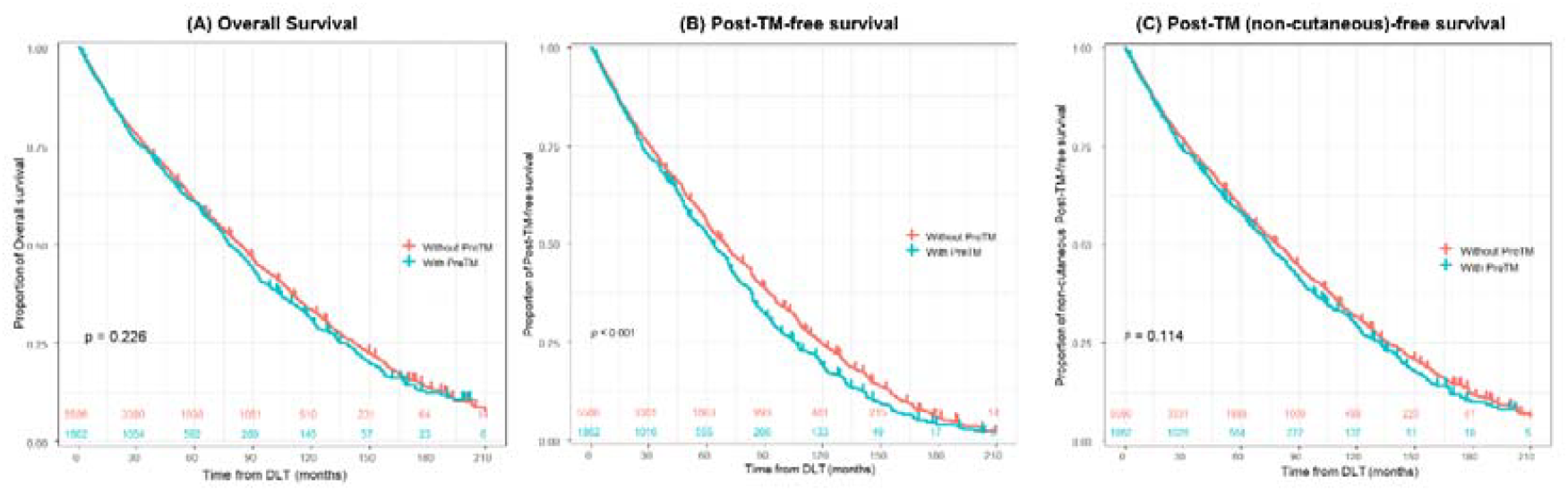
Clinical outcomes of the cohort after propensity-score matching according to the status of pre-transplant malignancy. Pre-TM, pre-transplant malignancy; Post-TM, post-transplant malignancy

Pre-TM was identified as a significant risk factor for both overall (hazard ratio [HR] 1.12, 95% confidence interval [CI] 1.04-1.21, p=0.004) and Post-TM-free survival (HR 1.20, 95% CI 1.12-1.28, p<0.001) across the entire cohort (Table 4). Nonetheless, when adjusted for confounding factors through propensity score matching, this risk notably decreased for overall survival (HR 1.06, 95% CI 0.98-1.16, p=0.163). Similarly, the impact on Post-TM was not statistically significant after skin cancers were excluded (HR 1.08, 95% CI 0.99-1.17, p=0.068) (Table 5).

### Recurrence of pre-transplant malignancy

When comparing the post-transplant recurrence of Pre-TM to the incidence of de novo malignancies of the same cancer type post-transplant, no statistical differences were observed in most types of cancer: breast (p=0.732), genitourinary (p=0.633), head and neck (p=0.147), hematologic (p=0.213), neurologic (p=1.000), respiratory (p=0.472), except for skin cancer (p<0.001) (Table 6).

## Discussion

This study is the first of its kind, analyzing the latest patient data to offer a comparative assessment of outcomes between DLT recipients with or without Pre-TM, and between the post-transplant recurrence of Pre-TM and the incidence of de novo Post-TM. Our findings reveal that, after controlling for confounding factors with propensity score matching, there is no significant difference in overall and Post-TM-free survival rates between DLT recipients, regardless of Pre-TM status. Moreover, the rate of Pre-TM recurrence post-transplant closely matches the rate of Post-TM of the same type occurring de novo, with the sole exception being skin cancer.

The notion that Pre-TM leads to worse clinical outcomes originates from studies on kidney transplant recipients^12^. However, the demographics and pre-transplant treatments of patients awaiting kidney transplants differ markedly from those of DLT recipients. Unlike end-stage renal disease patients who may receive hemodialysis, individuals with end-stage lung diseases lack similar bridging treatments, making it more challenging for them to wait out the recommended periods between cancer treatment due to their high-risk diseases and comorbidities. The impact of Pre-TM on the survival of DLT recipients may also vary from its impact on other organ transplants, given the elevated risk of graft failure associated with DLTs. Hence, there is a need to reassess the implications of Pre-TM in the context of DLT.

The higher incidence of Post-TM among DLT recipients compared to other organ transplants may stem from the high levels of immunosuppression required for DLT.^7^ This increased immunosuppression, necessitated by poorer allograft survival rates, is related with ischemia-reperfusion injury and infection, leading to early graft injury in DLT and activating alloantigen-specific T-cell expansion. The resultant inflammation, alongside the effects of humoral and innate immunity, yields a prognosis for graft function in DLT that differs significantly from that of other transplants.^4,7^ Moreover, DLT recipients’ increased susceptibility to viral infections, which are frequently associated with carcinogenesis, further heightens the risk of Post-TM. The use of calcineurin inhibitors, which is known to promote cancer progression more so than mTOR inhibitors, remains prevalent among DLT recipients, adding another layer of risk.^13,14^ Given these factors, DLT patients, particularly those with a Pre-TM, face an even greater likelihood of Post-TM due to both the intense immunosuppression and their potential genetic predisposition to cancer.

The types of Post-TM developed in the context of immunosuppression is distinct from the nature of cancer commonly seen in the general population.^15^ This distinction also extends to patients with Pre-TM, where, in our study, the recurrence of the primary cancer post-transplant was infrequent, with the notable exception of cutaneous malignancies. Such skin cancers are recognized for their elevated recurrence risk compared to other cancer types among kidney transplant recipients.^16^ Conversely, the risk of recurrence for non-cutaneous malignancies among DLT recipients appears to lower, a phenomenon that may be attributed to comprehensive surveillance prior to DLT or to differences in the biological processes of malignancy following the transplant. This observation implies that DLT does not necessarily increase the risk of recurrent solid organ tumors, emphasizing the importance of considering broader factors beyond cancer history in DLT eligibility, particularly for those with respiratory failure. This approach prevents excluding patients from life-saving DLT based on their cancer history, aligning with the goal of enhancing patient outcomes through customized clinical decisions.

Despite the higher risk of Post-TM for patients with Pre-TM, our study suggests that this does not directly translate to poorer overall survival rates. In the past, cancers in transplant recipients were often diagnosed at more advanced stages compared to those in the general population. This trend, however, has been shifting thanks to the implementation of comprehensive surveillance systems and the establishment of detailed guidelines aimed at early detection and treatment. The heightened awareness and vigilance for cancer among the Pre-TM group, coupled with the routine check-ups DLT recipients undergo, likely play a pivotal role in mitigating the adverse effects on clinical outcomes. Furthermore, the landscape of cancer treatments has evolved dramatically with the advent of immunotherapy and the adoption of multidisciplinary care approaches, fundamentally altering the outlook of patients with cancer and substantially prolonging survival. In our study, which analyzes the latest data reflecting advancements in treatment, we provide compelling evidence of the potential clinical benefits for patients with Pre-TM undergoing DLT. The introduction of enhanced cancer screening protocols and individualized immunosuppressive therapy for these high-risk patients further bolsters the possibility of achieving better outcomes.^17^

While the management and care of DLT recipients with Pre-TM have seen significant advancements, several considerations remain for identifying the most appropriate candidates for transplantation. Among these, determining the optimal wait-time during the no-evidence of disease (NED) period is vital, as it indirectly reflects the aggressiveness of cancers and informs the ideal timing for DLT. Additionally, devising strategies for the ongoing monitoring and screening for Post-TM in these high-risk patients is imperative. Incorporating age-specific risk factors could enable a more personalized approach to cancer screening approach, given the apparent correlation between age and Post-TM risk.^17^ The application of cell-free DNA monitoring offers a promising, minimally invasive method to detect cancer recurrence or the emergence of new malignancies.^18^ Exploring personalized immunotherapy options could further minimize the need for chronic immunosuppression, which is particularly crucial for transplant recipients with cancer risks.^19^ In this context, we are conducting the registration study (NCT 05671887)^20^ which aimed to evaluate the risk and benefits of DLT in patients with a recent history of malignancy or active, lung-limited cancer. The promising preliminary data from this trial are anticipated to provide a more in-depth understanding of Pre-TM’s impacts on transplant outcomes, paving the way for enhanced patient selection criteria and post-transplant care.^21^

This study faces several limitations that warrant careful consideration. Firstly, the classification of Pre- and Post-TM was based solely on the organ system, lacking detailed histologic, biologic, and staging information crucial for predicting patient’s prognosis. This approach could potentially lead to an overestimation of both the incidence of malignancies and the recurrence rates of primary cancers. Secondly, current and previous guidelines and literature likely resulted in the exclusion of many Pre-TM patients from DLT, meaning our study cohort predominantly consists of individuals with relatively early-stage or slow-growing tumors. Moreover, the inherent constraints of large database studies and retrospective reviews limit their direct applicability to the clinical setting. Factors not captured in the registry, yet vital for clinical prediction -- beyond what the Lung Allocation Score (LAS) can forecast -- highlight the need for a nuanced understanding of patient selection and prognosis.^22^ Lastly, the absence of detailed information on immunosuppression regimens prevents a comprehensive analysis of their impact. Given the patient-specific nature of immunosuppressant levels and vulnerability to infections, further institutional-level analysis is essential to elucidate the effects of immunosuppressive therapies.

In summary, despite observing a higher incidence of Post-TM, patients with Pre-TM showed overall survival rates after DLT that were comparable to those without Pre-TM. Interestingly, with the exception of skin cancers, the risk of developing Post-TM was not significantly higher in the Pre-TM group. Moreover, the likelihood of the primary cancer recurring post-transplant did not exceed that in patients without a history of malignancy. These findings indicate that transplantation remains a viable option for individuals with Pre-TM. However, managing immunosuppression after transplant demands a careful balance between the risk of organ rejection and the potential for post-transplant malignancy and mortality. This equilibrium is essential for optimizing outcomes and highlights the need for tailored post-transplant care strategies for patients with Pre-TM.

## List of Abbreviations

DLT: Double Lung Transplant
Pre-TM: Pre-transplant Malignancy
Post-TM: Post-transplant Malignancy
UNOS: United Network for Organ Sharing
COPD: Chronic Obstructive Pulmonary Disease
IPF: Idiopathic Pulmonary Fibrosis
CF: Cystic Fibrosis
ISHLT: International Society for Heart and Lung Transplantation
IQR: Interquartile Range
ECOG: Eastern Cooperative Oncology Group
NED: No Evidence of Disease
LAS: Lung Allocation Score

## Author contributions

**WW:** Conceptualization, Methodology, Data Curation, Formal analysis, Investigation, Software, Writing - Original Draft, Writing – Review and Editing. **HSK:** Conceptualization, Methodology, Data Curation, Formal analysis, Investigation, Software, Writing - Original Draft, Writing – Review and Editing. **AB:** Conceptualization, Methodology, Supervision, Project administration, Writing – Review and Editing. **YKC:** Conceptualization, Methodology, Validation, Supervision, Project administration, Writing – Review and Editing.

## Acknowledgements

None.

## Ethical Approval statement

Patient information reported in the registry is deidentified and publicly available. Thus, this research was determined to be exempt from review by our institution’s Institutional Review Board.

## Data Availability Statement

The data underlying this article will be shared by the corresponding author upon reasonable request.

