## Supplementary material for "Rethinking the Impact of Pretransplant Malignancy (Pre-TM) on Double Lung Transplantation (DLT) Eligibility: An Analysis of 23,291 DLT Recipients": Table

**Table 1. Clinical characteristics of DLT patients according to the status of pre-transplant malignancy**

| Clinical Variables | Overall | Pre-TM (-) | Pre-TM (+) |  |
| --- | --- | --- | --- | --- |
|  | N=23291 | N=21421 | N=1870 | p.value |
| Age | 58.0 [48.0, 64.0] | 57.0 [47.0, 64.0] | 63.0 [57.0, 67.0] | <0.001 |
| Sex |  |  |  | 0.028 |
| Male | 13582 (58.3) | 12446 (58.1) | 1136 (60.7) |  |
| Female | 9709 (41.7) | 8975 (41.9) | 734 (39.3) |  |
| Race |  |  |  | <0.001 |
| White | 18307 (78.6) | 16630 (77.6) | 1677 (89.7) |  |
| Black | 2325 (10.0) | 2243 (10.5) | 82 (4.4) |  |
| Hispanic | 1969 (8.5) | 1888 (8.8) | 81 (4.3) |  |
| Asian | 486 (2.1) | 469 (2.2) | 17 (0.9) |  |
| Others | 204 (0.9) | 191 (0.9) | 13 (0.7) |  |
| Comorbidities |  |  |  |  |
| Diabetes mellitus | 4479 (19.3) | 4192 (19.6) | 287 (15.4) | <0.001 |
| Smoking history | 12708 (55.4) | 11518 (54.6) | 1190 (64.0) | <0.001 |
| History of cardiac surgery | 700 (3.0) | 630 (2.9) | 70 (3.7) | 0.061 |
| History of dialysis | 99 (0.4) | 90 (0.4) | 9 (0.5) | 0.838 |
| Performance scale |  |  |  | 0.006 |
| ECOG 0-1 | 5016 (21.9) | 4566 (21.7) | 450 (24.5) |  |
| ECOG 2-3 | 17900 (78.1) | 16510 (78.3) | 1390 (75.5) |  |
| **Preop evaluation variables** |  |  |  |  |
| Lung Allocation Score | 41 [35, 56] | 42 [35, 56] | 41 [35, 53] | <0.001 |
| FEV1, % | 34 [21, 53] | 33 [21, 52] | 38 [22, 58] | <0.001 |
| FVC, % | 46 [35, 59] | 46 [35, 59] | 49 [37, 62] | <0.001 |
| Cardiac output, L/min | 5 [4, 6] | 5 [4, 6] | 5 [5, 6] | 0.011 |
| Mean PA pressure | 26 [21, 33] | 26 [21, 33] | 25 [20, 31] | <0.001 |
| Mean PCWP | 10 [7, 14] | 10 [7, 14] | 10 [7, 14] | 0.021 |
| **Seropositivity of virus** |  |  |  |  |
| CMV positive | 12612 (55.5) | 11650 (55.7) | 962 (53.1) | 0.033 |
| EBV positive | 19983 (85.8) | 18339 (90.5) | 1644 (92.8) | 0.002 |
| HBV positive | 323 (1.4) | 294 (1.4) | 29 (1.6) | 0.605 |
| HCV positive | 499 (2.2) | 456 (2.2) | 43 (2.4) | 0.658 |
| HIV positive | 50 (0.2) | 43 (0.2) | 7 (0.4) | 0.186 |
| **Transplant related factors** |  |  |  |  |
| Mean wait time, days | 51 [15, 169] | 52 [15, 172] | 43 [13, 137] | <0.001 |
| Ischemic time, hours | 6 [5, 7] | 6 [5, 7] | 6 [5, 7] | 0.104 |
| HLA mismatch | 21509 (99.9) | 19760 (99.9) | 1749 (99.9) | 1 |
| Number of HLA mismatched loci | 5 [4, 5] | 5 [4, 5] | 5 [4, 5] | 0.428 |
| Preoperative ECMO | 658 (2.8) | 626 (2.9) | 32 (1.7) | 0.003 |
| Preoperative MV | 2570 (11.0) | 2408 (11.2) | 162 (8.7) | 0.001 |
| Indication for DLT |  |  |  | <0.001 |
| Cystic fibrosis | 3017 (13.0) | 2968 (13.9) | 49 (2.6) |  |
| COPD | 5024 (21.6) | 4544 (21.2) | 480 (25.9) |  |
| IPF | 8580 (36.9) | 7742 (36.1) | 838 (45.2) |  |
| PPH | 736 (3.2) | 689 (3.2) | 47 (2.5) |  |
| Other | 5918 (25.4) | 5478 (25.6) | 440 (23.7) |  |
| **Clinical outcome** |  |  |  |  |
| Post-transplant malignancy | 4623 (19.8) | 4116 (19.2) | 507 (27.1) | <0.001 |
| Graft failure | 4224 (18.4) | 3967 (18.8) | 257 (13.9) | <0.001 |
| Graft failure due to AR | 202 (0.9) | 183 (0.9) | 19 (1.0) | 0.568 |
| Graft failure due to CR | 2357 (10.3) | 2220 (10.5) | 137 (7.4) | <0.001 |
| Mortality | 10400 (44.7) | 9555 (44.6) | 845 (45.2) | 0.645 |
| Cause of death |  |  |  |  |
| Cardiovascular | 623 (6.7) | 571 (6.7) | 52 (6.9) | 0.866 |
| Respiratory | 4339 (46.7) | 4041 (47.3) | 298 (39.6) | <0.001 |
| Malignancy | 1031 (11.1) | 907 (10.6) | 124 (16.5) | <0.001 |
| Others | 3304 (35.5) | 3026 (35.4) | 278 (37.0) | 0.415 |
| Follow-up time, months | 45.0 [20.0, 83.0] | 46.0 [21.0, 83.0] | 36.0 [14.0, 71.0] | <0.001 |

Data are presented as n(%) or median [interquartile range]

AR, acute rejection; CR, chronic rejection; CMV, cytomegalovirus; COPD, chronic obstructive pulmonary disease; DLT, double lung transplant; EBV, Epstein-Barr virus; ECMO, extracorporeal membrane oxygenation; ECOG, Eastern Cooperative Oncology Group; FEV1, Forced expiratory volume in the first second; FVC, Forced vital capacity; HBV, hepatitis B virus; HCV, hepatitis C virus; HIV, human immunodeficiency virus; HLA, human leukocyte antigens; IPF, idiopathic pulmonary fibrosis; PA, pulmonary artery; PCWP, pulmonary capillary wedge pressure; PPH, primary pulmonary hypertension; MV, mechanical ventilation;

**Table 2. Clinical characteristics of DLT patients according to the status of pre-transplant malignancy after propensity score matching**

| Clinical Variables | Pre-TM (-) | Pre-TM (+) |  |
| --- | --- | --- | --- |
|  | N=5588 | N=1862 | p-value |
| Age | 63.0 [57.0, 67.0] | 63.0 [57.0, 67.0] | 0.984 |
| Sex |  |  | 1 |
| Male | 3393 (60.7) | 1131 (60.7) |  |
| Female | 2193 (39.3) | 731 (39.3) |  |
| Race |  |  | 1 |
| White | 5019 (89.8) | 1673 (89.8) |  |
| Black | 243 (4.4) | 81 (4.4) |  |
| Hispanic | 237 (4.2) | 79 (4.2) |  |
| Asian | 51 (0.9) | 17 (0.9) |  |
| Others | 36 (0.6) | 12 (0.6) |  |
| Comorbidities |  |  |  |
| Diabetes mellitus | 960 (17.3) | 286 (15.4) | 0.078 |
| Smoking history | 3548 (64.3) | 1184 (64.0) | 0.817 |
| History of cardiac surgery | 208 (3.7) | 69 (3.7) | 1 |
| History of dialysis | 15 (0.3) | 9 (0.5) | 0.238 |
| Performance scale |  |  | 0.103 |
| ECOG 0-1 | 1241 (22.6) | 448 (24.5) |  |
| ECOG 2-3 | 4258 (77.4) | 1384 (75.5) |  |
| **Preop evaluation variables** |  |  |  |
| Lung Allocation Score | 41 [35, 54] | 41 [35, 53] | 0.553 |
| FEV1, % | 40 [23, 58] | 38 [22, 58] | 0.067 |
| FVC, % | 49 [38, 63] | 49 [37, 62] | 0.102 |
| **Seropositivity of virus** |  |  |  |
| CMV positive | 2993 (54.9) | 955 (53.0) | 0.164 |
| EBV positive | 4910 (92.3) | 1636 (92.7) | 0.587 |
| HBV positive | 74 (1.4) | 28 (1.5) | 0.637 |
| HCV positive | 125 (2.3) | 43 (2.4) | 0.885 |
| HIV positive | 9 (0.2) | 7 (0.4) | 0.140 |
| **Transplant related factors** |  |  |  |
| Mean wait time, days | 47.0 [13.0, 158.0] | 43.5 [13.0, 137.0] | 0.072 |
| Ischemic time, hours | 6 [5, 7] | 6.0 [5.0, 7.0] | 0.524 |
| HLA mismatch | 5199 (99.9) | 1742 (99.9) | 1 |
| Number of HLA mismatched loci | 5 [4, 5] | 5 [4, 5] | 0.115 |
| Preoperative ECMO | 85 (1.5) | 32 (1.7) | 0.628 |
| Preoperative MV | 466 (8.3) | 161 (8.6) | 0.718 |
| Indication for DLT |  |  | 0.105 |
| Cystic fibrosis | 318 (5.7) | 49 (2.7) |  |
| COPD | 1464 (26.2) | 480 (26.0) |  |
| IPF | 2507 (44.9) | 832 (45.1) |  |
| PPH | 1158 (20.7) | 438 (23.7) |  |
| Other | 139 (2.5) | 47 (2.5) |  |
| **Clinical outcome** |  |  |  |
| Post-transplant malignancy | 1276 (22.8) | 506 (27.2) | <0.001 |
| Graft failure | 922 (16.7) | 256 (13.9) | 0.005 |
| Graft failure due to AR | 32 (0.6) | 19 (1.0) | 0.064 |
| Graft failure due to CR | 488 (8.8) | 136 (7.4) | 0.054 |
| Mortality | 2612 (46.8) | 843 (45.3) | 0.277 |
| Cause of death |  |  |  |
| Cardiovascular | 175 (7.5) | 52 (6.9) | 0.645 |
| Respiratory | 995 (42.8) | 296 (39.5) | 0.118 |
| Malignancy | 281 (12.1) | 124 (16.5) | 0.002 |
| Others | 874 (37.6) | 278 (37.1) | 0.830 |
| Follow-up time, months | 40.0 [16.0, 75.0] | 36.0 [14.0, 71.0] | 0.001 |

Data are presented as n (%) or median [interquartile range]

AR, acute rejection; CR, chronic rejection; CMV, cytomegalovirus; COPD, chronic obstructive pulmonary disease; DLT, double lung transplant; EBV, Epstein-Barr virus; ECMO, extracorporeal membrane oxygenation; ECOG, Eastern Cooperative Oncology Group; FEV1, Forced expiratory volume in the first second; FVC, Forced vital capacity; HBV, hepatitis B virus; HCV, hepatitis C virus; HIV, human immunodeficiency virus; HLA, human leukocyte antigens; IPF, idiopathic pulmonary fibrosis; PA, pulmonary artery; PCWP, pulmonary capillary wedge pressure; PPH, primary pulmonary hypertension; MV, mechanical ventilation;

**Table 3. Cancer types of patients with pre-transplant malignancy**

| **Pretransplant malignancy** |  | **N=23291** |
| --- | --- | --- |
| **None** |  | **21421** |
| **Any pre-transplant malignancy** | | **1870** |
|  | Skin | 726 (39.0) |
|  | Breast | 197 (10.6) |
|  | Genitourinary | 191 (10.3) |
|  | Head and Neck | 50 (2.7) |
|  | Hematologic | 148 (7.9) |
|  | CNS origin | 6 (0.3) |
|  | Respiratory | 51 (2.7) |
|  | Multiple | 113 (6.1) |
|  | Unknown | 6 (0.3) |
|  | Others | 374 (20.1) |

**Table 4. Cox-proportional analysis of risk factors in the entire study cohort**

| Outcome | Overall Survival | | | Post-TM-Free survival | | | Non-cutaneous Post-TM-Free survival | | |
| --- | --- | --- | --- | --- | --- | --- | --- | --- | --- |
| Clinical Variables | HR | 95% CI | p-value | HR | 95% CI | p-value | HR | 95% CI | p-value |
| **Unadjusted Pre-TM (+)** | **1.20** | **1.12-1.29** | **<0.001** | **1.32** | **1.24-1.41** | **<0.001** | **1.21** | **1.13-1.29** | **<0.001** |
| *Multivariate model* |  |  |  |  |  |  |  |  |  |
| **Pre-TM (+)** | **1.12** | **1.04-1.21** | **0.004** | **1.20** | **1.12-1.28** | **<0.001** | **1.13** | **1.05-1.22** | **0.002** |
| Age (years) | 1.01 | 1.01-1.01 | <0.001 | 1.02 | 1.01-1.02 | <0.001 | 1.01 | 1.01-1.02 | <0.001 |
| Female | 0.98 | 0.93-1.02 | 0.262 | 0.90 | 0.86-0.94 | <0.001 | 0.97 | 0.93-1.01 | 0.132 |
| Race |  |  |  |  |  |  |  |  |  |
| Caucacian | Ref | Ref |  | Ref | Ref |  | Ref | Ref |  |
| Black | 1.07 | 1.00-1.15 | 0.068 | 0.91 | 0.85-0.98 | 0.007 | 1.07 | 1.00-1.15 | 0.062 |
| Hispanic | 0.96 | 0.88-1.05 | 0.373 | 0.87 | 0.81-0.94 | <0.001 | 0.95 | 0.88-1.04 | 0.273 |
| Asian | 0.77 | 0.64-0.92 | 0.004 | 0.67 | 0.56-0.79 | <0.001 | 0.76 | 0.64-0.90 | 0.002 |
| Others | 1.14 | 0.92-1.43 | 0.232 | 1.00 | 0.81-1.24 | 0.983 | 1.13 | 0.90-1.40 | 0.293 |
| Diabetes mellitus | 1.15 | 1.09-1.21 | <0.001 | 1.10 | 1.04-1.16 | <0.001 | 1.14 | 1.08-1.20 | <0.001 |
| Smoking history | 1.03 | 0.98-1.09 | 0.222 | 1.00 | 0.95-1.04 | 0.832 | 1.03 | 0.98-1.09 | 0.224 |
| ECOG 2 or 3 | 1.07 | 1.02-1.13 | 0.011 | 1.04 | 0.99-1.09 | 0.124 | 1.06 | 1.00-1.12 | 0.031 |
| Hemodialysis history | 1.52 | 1.14-2.02 | 0.005 | 1.44 | 1.09-1.90 | 0.010 | 1.47 | 1.10-1.96 | 0.009 |
| Pre-transplant MV | 1.11 | 1.03-1.20 | 0.006 | 1.11 | 1.04-1.19 | 0.003 | 1.12 | 1.04-1.21 | 0.002 |
| LAS | 1.00 | 1.00-1.00 | 0.001 | 1.00 | 1.00-1.00 | <0.001 | 1.00 | 1.00-1.00 | <0.001 |
| Ischemic time (hours) | 1.02 | 1.01-1.03 | <0.001 | 1.02 | 1.01-1.03 | <0.001 | 1.02 | 1.00-1.03 | 0.004 |
| Indication for DLT |  |  |  |  |  |  |  |  |  |
| Cystic Fibrosis | Ref | Ref |  | Ref | Ref |  | Ref | Ref |  |
| COPD | 1.12 | 1.00-1.24 | 0.044 | 1.06 | 0.96-1.17 | 0.283 | 1.09 | 0.98-1.22 | 0.095 |
| IPF | 1.05 | 0.96-1.16 | 0.293 | 1.01 | 0.92-1.10 | 0.894 | 1.02 | 0.93-1.12 | 0.665 |
| PPH | 1.13 | 0.98-1.31 | 0.143 | 1.07 | 0.93-1.23 | 0.352 | 1.10 | 0.95-1.27 | 0.234 |
| Others | 1.00 | 0.91-1.10 | 0.973 | 0.98 | 0.90-1.07 | 0.682 | 0.98 | 0.89-1.07 | 0.642 |

CI, confidence interval; COPD, chronic obstructive pulmonary disease; ECOG, Eastern Cooperative Oncology Group; HR, hazard ratio; IPF, idiopathic pulmonary fibrosis; LAS, lung allocation score; PPH, primary pulmonary hypertension; Post-TM, post-transplant malignancy; Pre-TM, Pre-transplant malignancy; MV, mechanical ventilation

**Table 5. Cox-proportional analysis of risk factors in the propensity-score matched study cohort**

| Outcome | Overall Survival | | | Post-TM-Free survival | | | Non-cutaneous Post-TM-Free survival | | |
| --- | --- | --- | --- | --- | --- | --- | --- | --- | --- |
| Clinical Variables | HR | 95% CI | p-value | HR | 95% CI | p-value | HR | 95% CI | p-value |
| **Unadjusted Pre-TM (+)** | **1.05** | **0.97-1.13** | **0.233** | **1.13** | **1.06-1.22** | **<0.001** | **1.06** | **0.99-1.15** | **0.122** |
| *Multivariate model* |  |  |  |  |  |  |  |  |  |
| **Pre-TM (+)** | **1.06** | **0.98-1.16** | **0.163** | **1.15** | **1.07-1.24** | **<0.001** | **1.08** | **0.99-1.17** | **0.068** |
| Age (years) | 1.02 | 1.02-1.03 | <0.001 | 1.03 | 1.02-1.03 | <0.001 | 1.02 | 1.02-1.03 | <0.001 |
| Female | 0.87 | 0.80-0.94 | <0.001 | 0.81 | 0.76-0.87 | <0.001 | 0.86 | 0.80-0.93 | <0.001 |
| Race |  |  |  |  |  |  |  |  |  |
| Caucacian | Ref | Ref |  | Ref | Ref |  | Ref | Ref |  |
| Black | 1.10 | 0.92-1.31 | 0.322 | 0.97 | 0.81-1.14 | 0.683 | 1.10 | 0.93-1.31 | 0.282 |
| Hispanic | 1.09 | 0.89-1.34 | 0.413 | 0.99 | 0.82-1.19 | 0.893 | 1.05 | 0.86-1.29 | 0.612 |
| Asian | 0.71 | 0.43-1.16 | 0.175 | 0.70 | 0.46-1.06 | 0.093 | 0.75 | 0.47-1.20 | 0.234 |
| Others | 1.23 | 0.78-1.94 | 0.373 | 1.04 | 0.68-1.60 | 0.852 | 1.23 | 0.79-1.91 | 0.363 |
| Diabetes mellitus | 1.08 | 0.98-1.20 | 0.103 | 1.05 | 0.96-1.14 | 0.332 | 1.08 | 0.98-1.19 | 0.114 |
| Smoking history | 1.08 | 0.99-1.18 | 0.086 | 1.01 | 0.94-1.10 | 0.723 | 1.08 | 0.99-1.18 | 0.089 |
| ECOG 2 or 3 | 1.08 | 0.98-1.18 | 0.122 | 1.05 | 0.97-1.14 | 0.253 | 1.07 | 0.98-1.17 | 0.153 |
| Hemodialysis history | 1.48 | 0.84-2.61 | 0.183 | 1.42 | 0.84-2.42 | 0.193 | 1.60 | 0.92-2.76 | 0.094 |
| Pre-transplant MV | 1.14 | 0.99-1.32 | 0.076 | 1.13 | 0.99-1.28 | 0.074 | 1.18 | 1.02-1.36 | 0.024 |
| LAS | 1.00 | 1.00-1.00 | 0.059 | 1.00 | 1.00-1.00 | 0.015 | 1.00 | 1.00-1.00 | 0.077 |
| Ischemic time (hours) | 1.01 | 0.99-1.03 | 0.322 | 1.00 | 0.96-1.04 | 0.232 | 1.01 | 0.99-1.03 | 0.293 |
| Indication for DLT |  |  |  |  |  |  |  |  |  |
| Cystic Fibrosis | Ref | Ref |  | Ref | Ref |  | Ref | Ref |  |
| COPD | 0.96 | 0.77-1.21 | 0.744 | 0.95 | 0.78-1.16 | 0.613 | 0.98 | 0.78-1.22 | 0.834 |
| IPF | 0.86 | 0.69-1.07 | 0.172 | 0.87 | 0.72-1.05 | 0.154 | 0.86 | 0.70-1.07 | 0.183 |
| PPH | 1.05 | 0.77-1.42 | 0.783 | 0.97 | 0.73-1.28 | 0.814 | 1.04 | 0.77-1.41 | 0.834 |
| Others | 0.83 | 0.67-1.03 | 0.089 | 0.86 | 0.71-1.04 | 0.124 | 0.83 | 0.68-1.03 | 0.095 |

CI, confidence interval; COPD, chronic obstructive pulmonary disease; ECOG, Eastern Cooperative Oncology Group; HR, hazard ratio; IPF, idiopathic pulmonary fibrosis; LAS, lung allocation score; PPH, primary pulmonary hypertension; Post-TM, post-transplant malignancy; Pre-TM, Pre-transplant malignancy; MV, mechanical ventilation

**Table 6. Incidence of same type post-transplant malignancy compared to patients without pre-transplant malignancy**

|  | **Post-transplant Malignancy incidence** | |  |
| --- | --- | --- | --- |
|  |  |  | p-value |
| **Pre-transplant malignancy** | Post Breast (+) | Post Breast (-) |  |
| No Pre-TM | 70 | 21351 | 0.732 |
| Breast Pre-TM (+) | 1 | 216 |  |
|  | Post GU (+) | Post GU (-) |  |
| No Pre-TM | 295 | 21126 | 0.633 |
| GU Pre-TM (+) | 4 | 225 |  |
|  | Post Head and Neck(+) | Post Head and Neck(-) |  |
| No Pre-TM | 88 | 21333 | 0.147 |
| Head and Neck Pre-TM (+) | 1 | 62 |  |
|  | Post Hema(+) | Post Hema(-) |  |
| No Pre-TM | 475 | 20946 | 0.213 |
| Hema Pre-TM (+) | 6 | 158 |  |
|  | Post Neuro (+) | Post Neuro (-) |  |
| No Pre-TM | 4 | 21417 | 1 |
| Neuro Pre-TM (+) | 0 | 7 |  |
|  | Post Lung cancer (+) | Post Lung Cancer (-) |  |
| No Pre-TM | 160 | 21261 | 0.472 |
| Lung Cancer Pre-TM (+) | 1 | 65 |  |
|  | Post Skin cancer (+) | Post Skin cancer (-) |  |
| No Pre-TM | 2858 | 18563 | <0.001 |
| Skin cancer Pre-TM (+) | 240 | 555 |  |

Breast, breast cancer; GU, genitourinary cancer; Head and Neck, head and neck cancer; Hema, hematologic malignancy; Neuro, central nervous system origin malignancy
